# Understanding Convergence Between Non-Hispanic Black and White COVID-19 Mortality: A County-Level Approach

**DOI:** 10.1101/2021.03.15.21253566

**Authors:** Ralph Lawton, Kevin Zheng, Daniel Zheng, Erich Huang

## Abstract

**Background:** Non-Hispanic Black populations have suffered greater per capita COVID-19 mortality at more than 1.5 times that of White populations. Previous work has established that, over time, rates of Black and White mortality have converged; however, some studies suggest that regional shifts in COVID-19 prevalence may play a role in the relative change between racial groups. This study’s objective was to investigate changes in Black and White COVID-19 mortality over time and uncover potential mechanisms driving these changes.

**Methods and Findings:** Using county-level COVID-19 mortality data stratified by race, we investigate the trajectory of non-Hispanic Black mortality, White mortality, and the Black/White per capita mortality ratio from June 2020–January 2021. Over this period, in the counties studied, cumulative mortality rose by 56.7% and 82.8% for Black and White populations respectively, resulting in a decrease in mortality ratio of 0.369 (23.8%). These trends persisted even when a county-level fixed-effects model was used to estimate changes over time within counties (controlling for all time-invariant county level characteristics and removing the effects of changes in regional distribution of COVID-19). Next, we leverage county-level variation over time in COVID-19 prevalence to show that the decline in the Black/White mortality ratio can be explained by changes in COVID-19 prevalence. Finally, we study heterogeneity in the time trend, finding that convergence occurs most significantly in younger populations, areas with less dense populations, and outside of the Northeast. Limitations include suppressed data in counties with fewer than 10 deaths in a racial category, and the use of provisional COVID-19 death data that may be incomplete.

**Conclusions:** The results of this study suggest that convergence in Black/White mortality is not driven by county-level characteristics or changes in the regional dispersion of COVID-19, but instead by changes within counties. Further, declines in the Black/White mortality ratio appear strongly linked to changes in COVID-19 prevalence, rather than a time-specific effect. Further studies on changes in exposure by race over time, or on the vulnerability of individuals who died at different points in the pandemic, may provide crucial insight on mechanisms and strategies to further reduce COVID-19 mortality disparities.

## 1. Introduction

From the early phases of the 2019 novel coronavirus disease (COVID-19) pandemic, numerous studies have documented disproportionately high numbers of COVID-19-attributed deaths in non-Hispanic Black populations, from here on referred to as Black, relative to those in non-Hispanic White populations, from here on referred to as White, in the US (Adhikari et al., 2020; Kirby, 2020; Laurencin & McClinton, 2020). Accordingly, racial disparities have remained a core part of national discussion. As of January 24th, 2021, the national per capita COVID-19 mortality for Black Americans is 1.47 times that of White Americans (Stokes et al., 2021). However, cross-sectional studies over different intervals during the pandemic suggest that there has been a recent convergence between White and Black mortality (Bassett et al., 2020; Gross et al., 2020; Webb Hooper et al., 2020).Estimates from April, May, and July 2020 document cumulative Black/White COVID-19 mortality ratios of 3.57, 2.37, and 1.97, respectively.

Although it is now well established that various socioeconomic and environmental factors are linked to racial disparities in COVID-19 outcomes, little work has yet been done in understanding the potential mechanisms for the observed convergence over time. Recent reports by Kumar et al. and Gold et al., two of the only longitudinal studies on COVID-19 mortality by race, investigated shifts in racial, ethnic, and geographic mortality from June to December 2020 and May to August 2020, respectively. Both papers documented relative declines in COVID-19 mortality among Black persons over this time frame, and Gold et al. found a shift in COVID-19 mortality from the Northeast and toward the South and West U.S. Census regions (Gold et al., 2020; Kumar et al., 2021). National press reporting, as well as these studies, attributed changes in racial mortality disparities in large part to the evolving geography of COVID-19 prevalence, due to regional differences in demographics (Zhou & Belluz, 2021). This theory suggests that, as COVID-19 prevalence shifted primarily from regions with proportionately larger Black populations to regions with proportionately smaller ones, national statistics may have recorded comparatively less Black mortality. However, while the West’s Black population is smaller than the Northeast’s as a percentage, the South’s is substantially larger, so it remains unclear whether regional changes in COVID-19 prevalence actually had a significant effect on racial mortality disparities. Other potential mechanisms that may explain the observed changes in racial mortality disparities include changes in exposure by race, ‘front-loading’ of mortality among more vulnerable population groups, or a variety of other potential factors. Research to further elucidate what mechanisms are significant is essential to fully understand COVID-19-related racial disparities and the myriad ways disease can disproportionately affect vulnerable populations.

These trends further highlight health disparities between Black and White people in the US. Generally, Black Americans have shorter lifespans relative to White Americans. The gap in life expectancy between Blacks and Whites is 3.6 years, but is projected to widen by 40% to more than 5 years due to the coronavirus pandemic (Andrasfay & Goldman, 2021). Black Americans also live a greater portion of their lives managing chronic conditions and disability due to differences in lifetime exposure to racism, stressors, and structural discrimination, all factors contributing to higher COVID-19 mortality rates among Black Americans (Lee & Hicken, 2016; Mays et al., 2007; Mitchell et al., 2021; Price-Haywood et al., 2020; Williams & Mohammed, 2009). Cross-sectional studies have also linked racial disparities in COVID-19 mortality to a variety of community-level factors including population density and urbanization. Black Americans often live in dense urban environments that can increase exposure to COVID-19 (Adhikari et al., 2020; Kirby, 2020; Stokes et al., 2021). Other studies have tied COVID-19 racial disparities to differences in access to care, economic status, chronic comorbidities, and “essential worker” categorization (Cheng et al., 2020; Paul et al., 2020; Wright et al., 2020). These studies primarily correlate the racial make-up of the population to total COVID-19 mortality, rather than disaggregated mortality data by race. Nonetheless, these findings suggest that disparities in COVID-19 mortality primarily reflect disproportionate COVID-19 prevalence in Black communities (Zelner et al., 2020).

While these cross-sectional studies have expanded our understanding of changing COVID-19 racial disparities, information about how racial, ethnic, geographic, and socioeconomic disparities have changed with regard to the observed convergence remains limited (Haleem et al., 2020). Previously identified factors such as population density, chronic comorbidities, and residential segregation have not changed significantly over the period of interest and are thus unlikely to have served as primary drivers for these observed changes in COVID-19 mortality between racial groups.

In this work, we use a county-level approach to further understand changes in COVID-19 mortality in Black and White populations over time. Using county-level data presents several key advantages, including the ability to investigate whether national trends are driven by changes within or between smaller areas. Furthermore, utilizing county-level variation in COVID-19 prevalence over time as well as county-level socioeconomic and population information enables us to address specific questions that are intractable at larger geographic areas.

We begin by examining county-level mortality in Black and White populations over time, documenting recent changes in mortality by race (including the extent to which observable control variables explain trends). Second, we use a county-level fixed-effects method to evaluate county-level changes in COVID-19 mortality by race. A variety of factors can make estimates of COVID-19 mortality time-trends difficult to interpret: notably, the changing geography of COVID-19 prevalence, the spread of COVID-19 to places with different demography, and different structural risk factors like population density, population-level chronic comorbidity rates, or housing supply, can complicate interpretation. Crucially, the fixed-effects approach enables us to estimate changes in these burdens *within* counties, such that our estimates are driven only by changes in COVID-19 mortality within small geographic areas. This approach allows us to control for all county-level characteristics, including many previously discussed structural risk factors, that can be considered fixed over this timeframe. Comparing fixed-effects estimates with traditional ordinary least squares estimates provide insight into the mechanisms that may underpin convergence. Third, we leverage county-level variation in COVID-19 prevalence to evaluate the extent to which time trends in disparities and mortality are attributed to changes in prevalence rather than a time effect. Finally, we investigate heterogeneity in these trends by region and county characteristics.

Understanding how disparities change throughout a pandemic is essential to understanding the differential burdens of a disease in real time. Our approach, which uses county-level fixed effects, develops a framework for investigating the origins of disparities in COVID-19 outcomes and provides insight into the potential mechanisms involved.

## 2. Methods and Data

### 2.1 Sample and Data

National and state COVID-19 mortality data were drawn from weekly provisional death counts reported to the National Vital Statistics System (NVSS) from May 6, 2020–January 27, 2021 (*Technical Notes*, 2020). National and state-level mortality data were utilized to generate visualizations of changing COVID-19 mortality by race and by state. County-level data was available from June 24, 2020–January 27, 2021. We restrict our analysis to the month of January as deaths after this period likely reflect exposures after substantial vaccination had occurred, potentially contaminating our estimates. COVID-19 deaths are identified using the *International Statistical Classification of Diseases and Related Health Problems* (ICD) code U07.1, where COVID-19 is an underlying or contributing cause of death. COVID-19 was the underlying cause for approximately 92% of deaths associated with COVID-19 (*COVID-19 Hospitalization and Death by Race/Ethnicity*, 2020). Provisional death counts are incomplete, and likely undercount true COVID-19 mortality. Calculations in October 2020 suggest that 94% of provisional death counts by age and race were complete as of that date (Andrasfay & Goldman, 2021). According to the NVSS, less than 0.7% of all recorded COVID-19 deaths are missing racial data (*Technical Notes*, 2020). To maintain patient confidentiality, counties with fewer than 10 deaths within a given racial group are suppressed by the NVSS. This represents a key limitation of the NVSS data. Suppression of data in counties with low numbers of total COVID-19 deaths means that our sample is not a random sample of American counties, though we do use some statistical methods to limit the effects of selection. Of the 579 counties represented in our dataset, 388 counties have mortality data for both non-Hispanic Black and White persons. These 388 counties represent 8,477 county-time observations and comprise our analytical sample. The counties in the analytical sample represent approximately 209 million Americans.

County-level demographic and population information are drawn from the 2018 5-year wave of the American Community Surveys (ACS). Combining county-level ACS and NVSS data, we construct our core outcomes. First, we generate per-capita estimates of COVID-19 mortality by race and ethnicity, dividing COVID-19-attributed deaths by total population within each racial group. For simplicity, we use COVID-19 deaths per 100,000 individuals, rather than the strict ratio of COVID-19 deaths to population. Our primary outcome is the natural logarithm of per capita COVID-19 mortality, as per capita mortality is non-normally distributed. Using the natural logarithm has the added advantage that coefficients over time can be interpreted as percent rates of change. To assess disparities, we construct ratios of COVID-19 mortality in Black populations divided by per capita COVID-19 mortality in White populations, building a metric for disparities between populations independent of total COVID-19 mortality.

Covariates for county-level household median income, age structure, population density, and occupation were drawn from the ACS data, and supplementary county-level data on COVID-19 testing was drawn from USAfacts.org, a standardized source for US government data.

### 2.2 Statistical Analysis

Ordinary least squares (OLS) regression is initially used to estimate time trends and differences between groups. We then use a county-level fixed effects model to estimate time trends and differences between groups *within* counties. Using this method, we are able to estimate the extent to which temporal changes in COVID-19 mortality and racial disparities are due to changes *within* compact geographic areas, rather than changes between regions and states with heterogeneous demography. The county fixed-effects method has the added benefit that results can be interpreted as conditional on all time-invariant county characteristics. In addition to location, factors such as county demography, population structure, racial and ethnic makeup, health infrastructure, and housing supply and quality can all be treated as fixed during this interval. Thus, the fixed-effects approach is crucial to generating interpretable estimates of the time-trend, especially as the changing geography of COVID-19 infection includes areas of differing demographic characteristics and structural factors, which can complicate estimates. Comparison between the fixed-effects estimates and OLS estimates may also shed insight into the extent to which these fixed characteristics may be related to the mechanisms involved.

In order to assess time effects in a semi-parametric fashion, indicator variables for each month are used as regression covariates. These models are extended to include covariates for the natural log of total mortality and three-week lagged positive COVID-19 tests, respectively, to identify the role of time trends versus changes in COVID-19 prevalence. Since COVID-19 prevalence is difficult to fully estimate, we use two different proxies for COVID-19 prevalence (total mortality and lagged testing) in order to check the robustness of different estimates. Finally, heterogeneity analyses for different time trends are conducted by generating interaction terms between the time indicators and covariates for regions, percentage of the population older than 85 years, and population density. Continuous regression covariates are de-meaned, so that regression intercepts can be interpreted for the mean county. In order to further investigate the role of age, state-level data are used to build parallel models by age and race. All standard errors reported are robust to heteroscedasticity. Analyses were conducted in Stata version 14.2.

## 3. Results

### 3.1 Sample Characteristics

The characteristics of counties included in the analytical sample are summarized in Table 1. The mean county in this sample is 49% male, 18% Black, 71% White, and has a population of approximately 539,000. As of January 27th, the observed counties experienced an average of 150.4 COVID-19-related deaths per 100,000 individuals. These data represent nearly two-thirds of the American population.

**Table 1:** Sample Characteristics. Summary statistics for counties represented in the analytical sample, including COVID-19 deaths per capita as of January 27th

|  | Mean | (Std. Error) |
| --- | --- | --- |
| <u>A. County Population</u> |  |  |
| Total Pop. | 538,644 | (40,446) |
| Pop./Sq. Mile | 1,533 | (246.6) |
| % Male | 48.82 | (0.0486) |
| % Over 85 | 1.936 | (0.0340) |
| % Black | 17.99 | (0.713) |
| % White | 70.86 | (0.735) |
| <u>B. County Economic Status</u> |  |  |
| HH Median Inc. | 61,345 | (901.1) |
| % Service Occupations | 17.92 | (0.142) |
| <u>C. Number of Counties by Region</u> |  |  |
| Northeast | 64 |  |
| Midwest | 81 |  |
| West | 29 |  |
| South | 214 |  |
| <u>D. COVID-19 Deaths</u> |  |  |
| COVID-19 Deaths/100k<br>population | 150.4 | (4.024) |
| Sample Size | 388 |  |
| (Standard Errors) in Parentheses |  |  |

### 3.2 National Trends in COVID-19 Mortality by Race

While Black populations have cumulatively suffered far greater per capita mortality than White populations, in recent months the cumulative ratio of deaths in Black populations versus White populations has declined significantly, driven by a convergence in mortality between these groups (Fig. 1a). Since November, weekly per capita mortality in White populations has surpassed that of Black populations (Fig. 1b). Although COVID-19 mortality in Black persons was mostly consistent from October to November, COVID-19 mortality in White persons grew significantly, and by January reached an all-time high. These results parallel similar convergence in COVID-19 prevalence by race (Appendix Fig. 1a-b).

**Figure 1:**
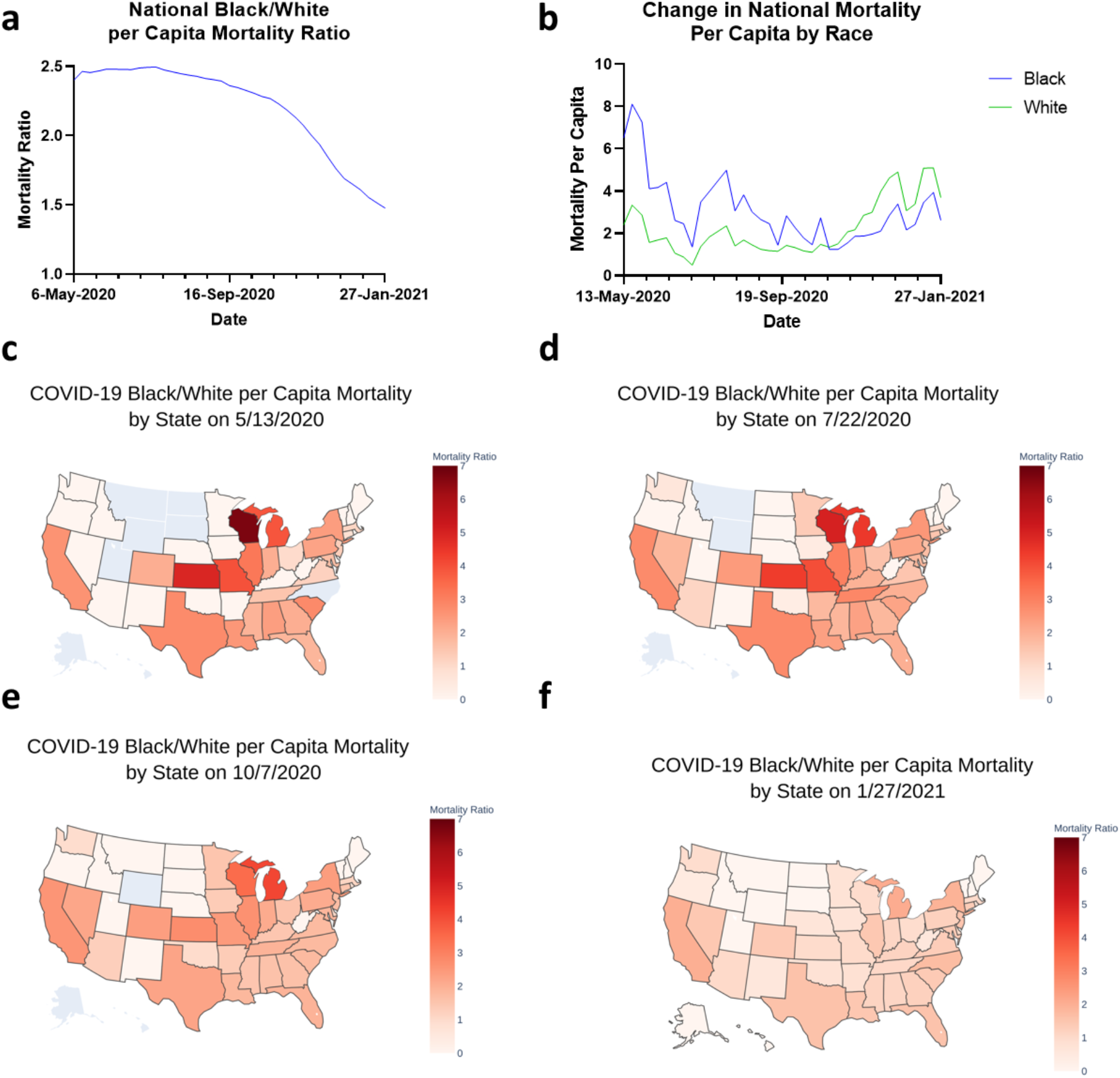
National Trends in COVID-19 Mortality by Race. (**a**)Weekly Black/White Mortality Ratio per Capita was calculated by date and subsequently plotted from 5/6/2020 to 1/27/2021. (**b**) Weekly Change in COVID-19 Mortality by race was calculated by date and subsequently plotted from 5/13/2020 to 1/27/2021. US maps were generated to visualize Black/White Mortality Ratio per Capita by state on (**c**) 5/13/2020, (**d**) 7/22/2020, (**e**) 10/7/2020, and (**f**) 1/27/2021.

Fig. 1c-f maps the ratio of COVID-19 mortality in Black versus White populations per capita by state in May, June, October, and January. Racial differences in COVID-19 mortality varied substantially across states in May. For instance, COVID-19 mortality was more than six times greater in the Black population relative to the White population in Wisconsin, and approximately three times greater in Kansas, Missouri, Michigan, and Illinois. However, COVID-19 mortality for Black and White populations was similar in Oregon, Washington, and Arizona. Nationally, the ratio of COVID-19 mortality between Black and White populations has declined over time. By January, the mortality ratio in nearly all states was less than three.

### 3.3 County-level Results

Table 2 provides information about the magnitude and direction of county-level COVID-19 mortality trends for Black persons [Column 1-2] (natural-log outcomes interpreted as percent change), White persons [Column 3-4], as well as the mortality ratio of the Black to White population from June 2020 to January 2021 [Column 5-6]. For each outcome, the left column presents the coefficients for time (months) from the regression model without control variables, and the right column presents the regression coefficients for time controlling for county-level characteristics.

**Table 2:** County-level Mortality by Race Over Time. County-level regressions for non-Hispanic Black and White mortality, as well as the mortality ratio with month indicator variables. Left column for each outcome reports time effects without covariates, and right column reports outcomes controlled for by de-meaned county-level characteristics.

| Outcome<br>Controls Applied? | [1]<br>ln(Black COVID-19<br>Mortality per capita) | [2]<br>ln(Black COVID-19<br>Mortality per capita) | [3]<br>ln(White COVID-19<br>Mortality per capita) | [4]<br>ln(White COVID-19<br>Mortality per capita) | [5]<br>Black/White per capita<br>COVID-19 Mortality | [6]<br>Black/White per capita<br>COVID-19 Mortality |
| --- | --- | --- | --- | --- | --- | --- |
|  | No | Yes | No | Yes | No | Yes |
| July | 0.0127<br>[0.141] | 0.0390<br>[0.510] | 0.00653<br>[0.0803] | 0.0291<br>[0.464] | -0.00781<br>[-0.0881] | -0.00220<br>[-0.0258] |
| August | 0.0869<br>[0.976] | 0.149<br>[1.986] | 0.0714<br>[0.896] | 0.129<br>[2.101] | -0.0121<br>[-0.138] | -0.00636<br>[-0.0753] |
| September | 0.189<br>[2.176] | 0.272<br>[3.703] | 0.190<br>[2.451] | 0.271<br>[4.496] | -0.0508<br>[-0.594] | -0.0515<br>[-0.624] |
| October | 0.282<br>[3.251] | 0.376<br>[5.111] | 0.315<br>[4.057] | 0.404<br>[6.721] | -0.103<br>[-1.201] | -0.101<br>[-1.223] |
| November | 0.343<br>[3.991] | 0.440<br>[6.040] | 0.423<br>[5.492] | 0.514<br>[8.609] | -0.160<br>[-1.891] | -0.158<br>[-1.933] |
| December | 0.451<br>[5.272] | 0.551<br>[7.588] | 0.631<br>[8.242] | 0.725<br>[12.17] | -0.269<br>[-3.168] | -0.273<br>[-3.318] |
| January | 0.567<br>[6.625] | 0.670<br>[9.221] | 0.828<br>[10.81] | 0.926<br>[15.52] | -0.369<br>[-4.349] | -0.378<br>[-4.597] |
| Constant | 4.341<br>[51.57] | 4.136<br>[56.76] | 4.034<br>[53.62] | 3.779<br>[63.27] | 1.548<br>[18.78] | 1.687<br>[20.48] |
| Observations | 8,477 | 8,477 | 8,477 | 8,477 | 8,477 | 8,477 |
| R-Squared | 0.061 | 0.306 | 0.154 | 0.497 | 0.025 | 0.117 |
| County FE? | NO | NO | NO | NO | NO | NO |

[T-statistics] in Brackets
June not shown
Controlled columns include de-meaned covariates for % of a County that is Black, % of people over 85, % male, median household income, population density, % in a service occupation, and US Census region indicators

Both White and Black populations have experienced a substantial increase in county-level total mortality over time. In June, the mortality for Black persons was greater than that for White persons; however, the increase in mortality has been much greater among White persons compared with Black persons from September onward. Relative to June, COVID-19 mortality increased by 1.3% by July, 18.9% by September, 45.1% by December, and 56.7% by January [Column 1] for Black persons; and 0.6% by July, 19% by September, 63.1% by December, and 82.8% by January for White persons [Column 3]. As a result of the greater increase in COVID-19 mortality among White populations, the Black/White mortality ratio has declined over time. Furthermore, these trends held even when controlled for by county characteristics [Columns 5-6].

Table 3 presents results from the fixed-effects models in which all observed and unobserved, time-invariant, county-level characteristics are taken into account. Fixed-effects estimates show similar time trends of COVID-19 mortality within counties. Columns 1 and 2 further support that total COVID-19 mortality has increased for both racial groups; however, greater increases in White mortality since September has resulted in a closing racial gap in recent months. By January 2021, the cumulative Black/White COVID mortality ratio had dropped by 0.343 (22.5%) compared to June [Column 3]. Consistency between the OLS regression and county fixed-effects models suggests that the shifting COVID mortality burden is not driven by changes in the regional distribution of COVID mortality, but rather by changes within counties. Our analysis reported similar results when a single fixed-effect model was used to estimate mortality trends for both Black and White populations together, compared to estimating the trend separately (Appendix 1).

**Table 3:** Fixed Effects Result at County Level. County-level fixed-effects regressions for non-Hispanic Black and White mortality, as well as the mortality ratio with month indicator variables. Columns 4 and 5 add covariates for COVID-19 disease prevalence at the county level.

| Outcome | [1]<br>ln(Black<br>COVID-19<br>Mortality<br>per capita) | [2]<br>ln(White<br>COVID-19<br>Mortality<br>per capita) | [3]<br>Black/White<br>per capita<br>COVID-19<br>Mortality | [4]<br>Black/White<br>per capita<br>COVID-19<br>Mortality | [5]<br>Black/White<br>per capita<br>COVID-19<br>Mortality |
| --- | --- | --- | --- | --- | --- |
| ln(Total Mort. Per Cap.) |  |  |  | -0.239<br>[-6.785] |  |
| ln(Lag Pos. Tests Per Cap.) |  |  |  |  | -0.224<br>[-7.117] |
| July | 0.0734<br>[4.229] | 0.0533<br>[2.899] | 0.0193<br>[1.530] | 0.0359<br>[2.877] | 0.0559<br>[4.519] |
| August | 0.209<br>[6.970] | 0.188<br>[5.775] | 0.0129<br>[0.574] | 0.0649<br>[2.914] | 0.120<br>[5.483] |
| September | 0.355<br>[9.948] | 0.355<br>[9.197] | -0.0240<br>[-0.967] | 0.0664<br>[2.533] | 0.147<br>[5.690] |
| October | 0.462<br>[11.98] | 0.487<br>[11.72] | -0.0678<br>[-2.541] | 0.0524<br>[1.848] | 0.148<br>[5.553] |
| November | 0.525<br>[13.26] | 0.587<br>[13.67] | -0.122<br>[-4.389] | 0.0184<br>[0.624] | 0.134<br>[4.755] |
| December | 0.647<br>[15.67] | 0.799<br>[17.58] | -0.244<br>[-7.953] | -0.0599<br>[-1.792] | 0.0847<br>[2.607] |
| January | 0.782<br>[18.25] | 1.012<br>[21.27] | -0.343<br>[-10.48] | -0.113<br>[-2.914] | 0.0715<br>[1.710] |
| Constant | 4.171<br>[118.8] | 3.882<br>[102.1] | 1.519<br>[61.14] | 1.390<br>[51.76] | 1.267<br>[46.55] |
| Observations | 8,477 | 8,477 | 8,477 | 8,477 | 8,477 |
| R-Squared | 0.566 | 0.641 | 0.335 | 0.389 | 0.400 |
| County FE? | YES | YES | YES | YES | YES |
| Number of Counties | 388 | 388 | 388 | 388 | 388 |

[T-statistics] in Brackets
June not shown
ln(Total Mort.) and ln(Lag Pos.) de-meanned

Columns 4 and 5 extend the models to include covariates to study the role of COVID-19 prevalence on the Black/White mortality ratio. Total per capita mortality and lagged positive per capita tests are both used to assess prevalence. Controlling for total mortality per capita and positive tests per capita nearly fully attenuates the downward trend in racial difference. In comparison, the January coefficient suggests a comparatively small time-specific effect. Overall, the coefficients approach 0, indicating no change over time. Thus, county-level variation in infection prevalence seems to act as the primary driver of time-based declines in Black/White COVID-19 mortality ratio. However, some small but statistically significant effects persist, such as in January, suggesting that the Black/White mortality ratio declined slightly more than expected during the January surge.

### 3.3 Heterogeneity by Age, Population Density, and Region

Assessment of heterogeneity in the time trends by region, age groups, and population density revealed that the Black/White COVID-19 mortality ratio has declined across all three age groups (0-64, 65-84, and ≥85 years) since June 2020; however, this gap has converged the most among persons aged less than 65 years. In June, the Black/White COVID-19 mortality gap was greatest among those under 65 years of age, with Black persons nearly six times more likely to die from COVID-19 than their White counterparts. By the end of January, the Black/White COVID-19 mortality ratio had declined by 1.99 from 4.75 in June, driven by rapid increases in COVID-19 mortality among young and middle-aged White patients.

Additionally, a significant negative interaction was observed between time and population density from September 2020 onward, with regard to cumulative Black and White COVID-19 mortality, indicating that counties with higher population density experienced relatively smaller increases in mortality. Conversely, regarding the racial gap, the positive interaction between time and population density suggests that counties with higher population density experienced less cumulative convergence between Black and White COVID-19 mortality since September.

Analysis by region revealed large geographic differences (Appendix 4).Compared to the time trends for the Northeast and Midwest, COVID-19 mortality increased more rapidly in the West and the South, for both Black and White populations. Similar to the national time trends shown in Table 3, COVID-19 mortality increased more among White persons in the Midwest, the West, and the South, resulting in larger declines in the racial gap over time. For example, among White persons living in the Midwest, total COVID mortality in January 2021 increased 125% relative to June 2020, while mortality among Black persons increased 85.5%. The considerable reduction in the Black/White COVID-19 mortality ratio in December reflects this increase in COVID-19 mortality among White persons in the Midwest. In the Northeast, increases in COVID-19 mortality have been comparatively similar for Black and White populations, resulting in an attenuated decline in the Black/White COVID-19 mortality ratio.

In order to assess whether the regional trends were driven by the region’s different stages of the pandemic (as suggested by the differing intercepts in June), prevalence covariates were added to an interacted model with regional coefficients (Appendix 5). This additional analysis revealed that differences between regions at each time point approach zero when accounting for COVID-19 prevalence, suggesting that differences in the Black/White mortality ratio by region primarily reflect different stages of the pandemic rather than differences in inherent regional characteristics.

## 4. Discussion

Our study set out to understand changes over time in the burdens of COVID-19 mortality in non-Hispanic Black and White populations, and the ratio between the two. This study makes three key contributions to the literature.

First, we investigate these trends over a comparatively long time period, from June 2020 through January 2021, during which an additional surge in cases occurred. Our results show that the national convergence between Black and White COVID-19 mortality outcomes was primarily driven by trends within counties. Previous work has suggested that changes in the regional dispersion of COVID-19 between countries or regions was driving national changes in the relationship between Black and White mortality (Gold et al., 2020; Kumar et al., 2021). However, our fixed-effects approach reveals that the driving force for the observed mortality convergence is occurring *within* counties. We find that even controlling for all county-specific time-invariant characteristics, these time trends persist.

Second, we show that the observed time trends can be almost entirely accounted for by the relationship between COVID-19 prevalence and the Black/White mortality ratio. Because COVID-19 cases rose over time, time and cumulative COVID-19 mortality are highly correlated. However, variations in COVID-19 prevalence at the county level enable us to separate the effects of time and prevalence, allowing us to identify a strong negative relationship between changes in COVID-19 prevalence and the Black/White mortality ratio that nearly fully accounts for the observed trends over time. This has important implications for understanding observed disparities in COVID-19 mortality, as linked to the trajectory of disease prevalence rather than specific activity at certain timepoints.

Third, we document heterogeneity in the time trends observed. We find that mortality in Black and White populations grew the most among individuals under 65 years of age. We also show that in younger populations, the Black/White mortality ratio fell the most. In addition, we find that places with higher population density experienced less COVID-19 mortality increases, and less convergence between Black and White outcomes. Finally, we show geographic variation in the time trend. While the overall pattern is similar between census regions, the Northeast saw smaller increases in COVID mortality over the specified interval, as well as less convergence between outcomes in Black and White populations. However, the regional variation can be nearly fully accounted for by changes in COVID-19 prevalence, suggesting that regions were at different stages of the pandemic, and providing further evidence for the core role of prevalence in driving mortality disparities rather than time.

Several potential mechanisms could explain the convergence over time between Black and White mortality, including changes in the regional prevalence of disease, exposure between populations, or vulnerability of individuals affected by COVID-19 over time. Our results suggest that the mechanism driving convergence is primarily affecting mortality within the county level, and is potentially linked to changes in cumulative COVID-19 prevalence rather than time. Given prior evidence that conditional on hospitalization, mortality among Black and White COVID-19 patients is similar, we discuss potential mechanisms upstream of hospitalization (Goyal et al., 2020; Krishnamoorthy et al., 2021; Moore, 2020; Renelus et al., 2020).

Regarding the role of regional prevalence, contrary to previous findings, our results suggest that regional changes in prevalence of COVID-19 are not the primary driver of convergence in racial mortality. Using the county fixed-effects approach, we show that the trends observed nationally are primarily occurring within, rather than between, counties. Furthermore, our results suggest that the primary mechanisms involved are taking place within smaller geographic areas, and that shifts in mortality trends are not accounted for by time-invariant county characteristics (such as population density or demographic composition).

While exposure between populations is a difficult mechanism to assess, particularly due to changing COVID-19 testing availability, the convergence over time between national per capita positive tests in Black and White populations parallels our core results (Appendix Figure 1). Changes over time in exposure to COVID-19 between the two populations is likely one of the driving forces in the observed mortality trends. The relationship between changes in exposure and changes in mortality is consistent with our findings on the effects of population density. Counties with dense populations, including cities, experienced more significant COVID-19 exposures earlier in the pandemic, and experienced less percent change in mortality later on. Furthermore, our results on the relationship between population density and mortality are consistent with evidence that areas of greater population density had both more effective and more persistent social distancing over time (Garnier et al., 2020). A potential behavioral mechanism may also be at play, and would be consistent with our results linking COVID-19 prevalence and mortality. Black populations faced much greater COVID-19 mortality earlier in the pandemic, and a combination of public health outreach and community exposure to the effects of the pandemic may have led to increased behavioral avoidance of COVID-19 later on.

Another potential mechanism includes temporal differences in mortality among vulnerable populations. It is possible that vulnerable groups may have had less control over exposure, particularly earlier in the pandemic, resulting in disproportionate COVID-19 infections and deaths early on, “front-loading” mortality in vulnerable groups. A wealth of literature has shown how systematic forces place Black populations at higher risk of exposure, including prevalence of “essential worker” classifications (Zelner et al., 2020). Furthermore, pre-existing conditions that could increase an individual’s likelihood of dying due to COVID-19 are more prevalent in Black populations. A combination of these health and socioeconomic factors may have resulted in increased vulnerability of Black populations relative to White populations early in the pandemic, contributing to the observed time trends. Prior evidence from persons with kidney failure is consistent with this theory: amongst this at-risk population, Black mortality exceeded White mortality by the largest margin early on in the pandemic (Kim et al., 2021). This mechanism would also be consistent with the relationship between the Black/White mortality ratio and changes in COVID-19 prevalence.

## 5. Limitations

This study has several limitations. We used provisional COVID-19 mortality data in which counties may have had differential delays in reporting deaths. Further, provisional death counts are incomplete, and so our study likely underestimates the cumulative number of deaths for any given interval. Additionally, due to confidentiality concerns, only counties with more than 10 deaths for both Black and White individuals are included. Although the counties used in the study comprise the majority of the U.S. population, they do not constitute a random subset of U.S. counties, and tend to have greater absolute populations and more COVID-19 deaths per capita than the average U.S. county. Therefore, while using the county fixed-effects approach controls for all fixed factors related to a county’s inclusion in our data, mitigating the effects of selection, our results may not generalize to counties not included in the sample. A third limitation is the lack of county-level data disaggregated by both age and race. Because our estimates are not age-adjusted, our findings likely understate the true difference in outcomes between Black and White populations. In addition, despite the use of a county fixed-effects framework, there may still be confounding factors that remain unaccounted for.

## 6. Conclusions

Between June 2020 and January 2021, per capita COVID-19 mortality increased more substantially for White populations than for Black populations, resulting in a declining Black/White COVID-19 mortality ratio. These effects persist even when controlling for all county-specific time-invariant factors, and are not driven by changes in the regional presence of COVID-19 prevalence. The observed convergence in Black/White COVID-19 mortality ratio occurred faster among younger populations and in counties with less dense populations, and slower in the Northeast where COVID-19 mortality grew less over the studied interval. Nearly all of the observed trends over time, including regional variation in the time trends, can be explained by changes in cumulative COVID-19 prevalence, suggesting that the primary driver of convergence between Black and White mortality outcomes is not time-specific, but rather due to the trajectory of infection prevalence.

Our findings suggest that the mechanisms that drive differences in mortality over time in Black and White populations are both occurring primarily within small geographic areas and are linked to changes in COVID-19 prevalence rather than time. Further research into potential mechanisms consistent with these criteria, such as changes in exposure and behavioral responses to prior mortality, or potential ‘front-loading’ of more-vulnerable patients, would be of high value. In particular, longitudinal studies of health behavior and of the characteristics of individuals who died earlier versus later in the pandemic may provide insight into the way vulnerability shapes mortality outcomes over the course of a long pandemic. This type of research will be important for fully understanding COVID-19-related mortality disparities, and may even shed light on some of the mechanisms driving health disparities broadly.

## Supporting information

County-Level Data Over Time

ICMJE Conflict of Interest Form

## Data Availability

All data is available in the manuscript or the supplemental materials. Data and materials will be fully available upon reasonable request.

## Acknowledgements

The authors thank D. Thomas, E. Frankenberg., J.S. Smith, Y. Zhang, K. Kadakia, and J. McCall for helpful discussion and thoughtful feedback. The authors thank J.Gold, F. Ahmad, and L. Rossen for providing COVID-19 mortality data by race. Funding: This work was supported by UL1TR002553 (E.H.).

## Author Contributions

R.L., K.Z., and E.H. conceived of the study. R.L. and D.Z. generated data constructs. R.L., K.Z., and D.Z analyzed the data. R.L., K.Z., D.Z., and E.H. wrote the paper. All authors discussed the results and reviewed the manuscript.

## Competing interests

E.S.H. is a non-executive Founder of Stratus Medicine, kēlaHealth, and MedBlue Data.

## Appendix

**Appendix 1:**
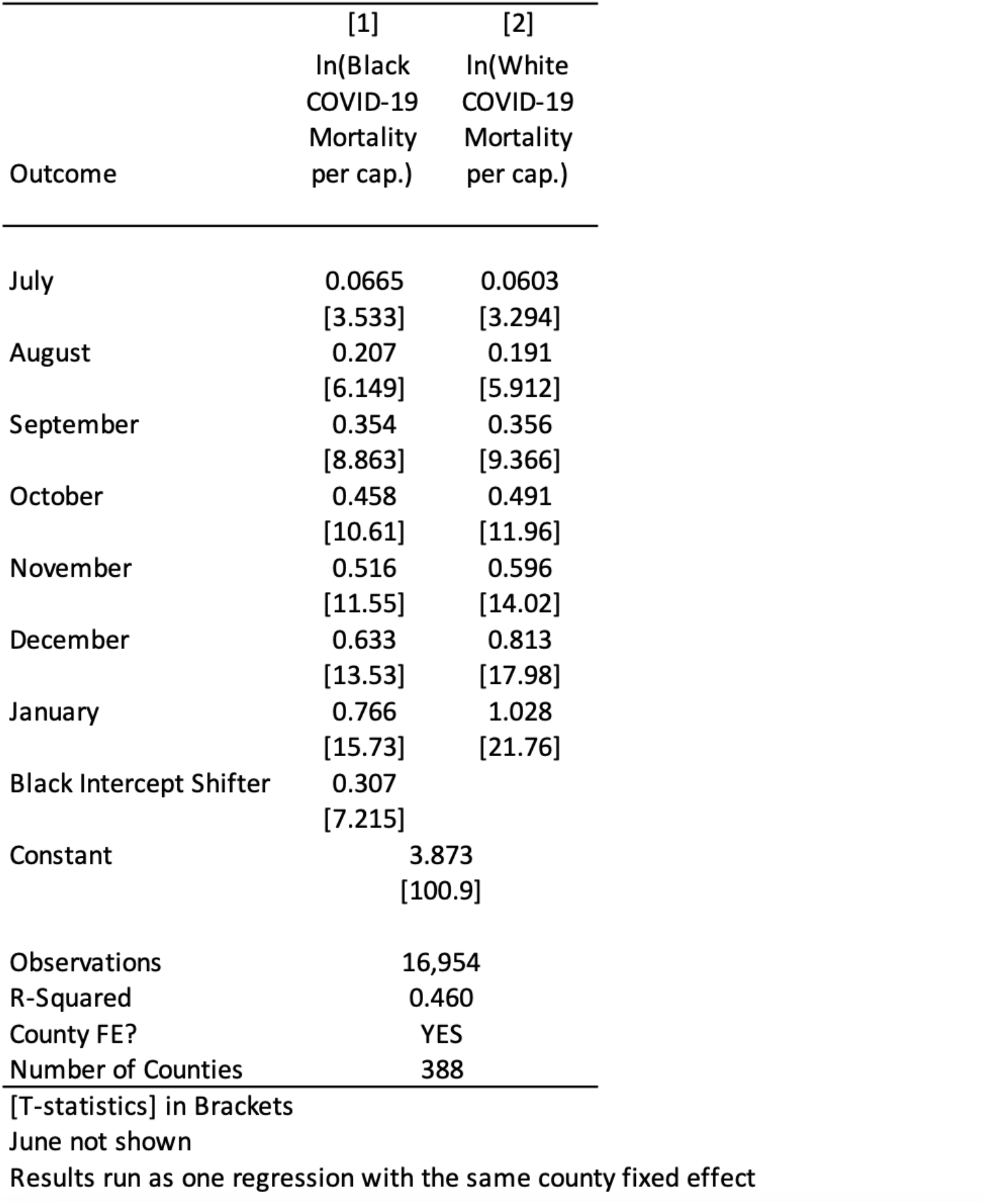
Core Fixed Effects Mortality results with same FE for both race groups. Black and White mortality over time estimated together utilizing a single fixed effect.

**Appendix 2:** Age Trends. County-level fixed-effects outcomes for non-Hispanic Black and White per capita mortality, and the mortality ratio, stratified by age category and utilizing month indicator variables over time.

| Age Group | [1] | [2] | [3] | [4] | [5] | [6] | [7] | [8] | [9] |
| --- | --- | --- | --- | --- | --- | --- | --- | --- | --- |
|  | Age<65 |  |  | Age 65-84 |  |  | Age 85+ |  |  |
| Outcome | ln(Black COVID-19 Mortality per cap.) | ln(White COVID-19 Mortality per cap.) | Black/White per capita COVID-19 Mortality | ln(Black COVID-19 Mortality per cap.) | ln(White COVID-19 Mortality per cap.) | Black/White per capita COVID-19 Mortality | ln(Black COVID-19 Mortality per cap.) | ln(White COVID-19 Mortality per cap.) | Black/White per capita COVID-19 Mortality |
| July | 0.571<br>[10.65] | 0.520<br>[10.18] | 0.0181<br>[0.109] | 0.537<br>[9.548] | 0.591<br>[16.73] | -0.00625<br>[-0.0358] | 0.570<br>[13.22] | 0.641<br>[17.54] | -0.0606<br>[-0.668] |
| August | 0.942<br>[11.54] | 0.951<br>[11.05] | -0.300<br>[-1.485] | 0.841<br>[11.77] | 0.965<br>[13.72] | -0.328<br>[-1.305] | 0.833<br>[11.42] | 0.980<br>[14.50] | -0.205<br>[-1.516] |
| September | 1.133<br>[10.89] | 1.234<br>[11.39] | -0.655<br>[-2.654] | 1.037<br>[11.94] | 1.211<br>[12.84] | -0.553<br>[-1.789] | 1.007<br>[10.78] | 1.204<br>[13.39] | -0.351<br>[-1.855] |
| October | 1.270<br>[11.19] | 1.456<br>[12.04] | -0.977<br>[-3.165] | 1.156<br>[11.94] | 1.404<br>[12.69] | -0.824<br>[-2.205] | 1.113<br>[10.71] | 1.380<br>[13.27] | -0.506<br>[-2.009] |
| November | 1.373<br>[11.77] | 1.629<br>[12.74] | -1.242<br>[-3.582] | 1.233<br>[12.17] | 1.590<br>[12.88] | -1.179<br>[-2.602] | 1.196<br>[11.20] | 1.563<br>[13.42] | -0.714<br>[-2.378] |
| December | 1.515<br>[12.63] | 1.909<br>[14.05] | -1.684<br>[-4.392] | 1.384<br>[13.11] | 1.877<br>[13.68] | -1.576<br>[-3.203] | 1.328<br>[12.04] | 1.841<br>[13.94] | -0.945<br>[-2.829] |
| January | 1.662<br>[13.34] | 2.168<br>[15.24] | -1.990<br>[-5.038] | 1.534<br>[13.84] | 2.128<br>[14.69] | -1.821<br>[-3.644] | 1.482<br>[13.06] | 2.087<br>[14.90] | -1.079<br>[-3.157] |
| Constant | 1.928<br>[23.32] | 0.438<br>[5.217] | 4.752<br>[20.41] | 4.923<br>[69.37] | 3.550<br>[45.29] | 4.066<br>[13.60] | 6.181<br>[81.90] | 5.302<br>[71.18] | 2.410<br>[12.22] |
| Observations | 1,281 | 1,343 | 1,242 | 1,324 | 1,401 | 1,322 | 1,265 | 1,400 | 1,263 |
| R-Squared | 0.727 | 0.766 | 0.339 | 0.718 | 0.761 | 0.267 | 0.701 | 0.750 | 0.218 |
| Number of States | 37 | 37 | 37 | 37 | 37 | 37 | 37 | 37 | 37 |
| County FE? | YES | YES | YES | YES | YES | YES | YES | YES | YES |

[T-statistics] in Brackets
June group not shown

**Appendix 3:**
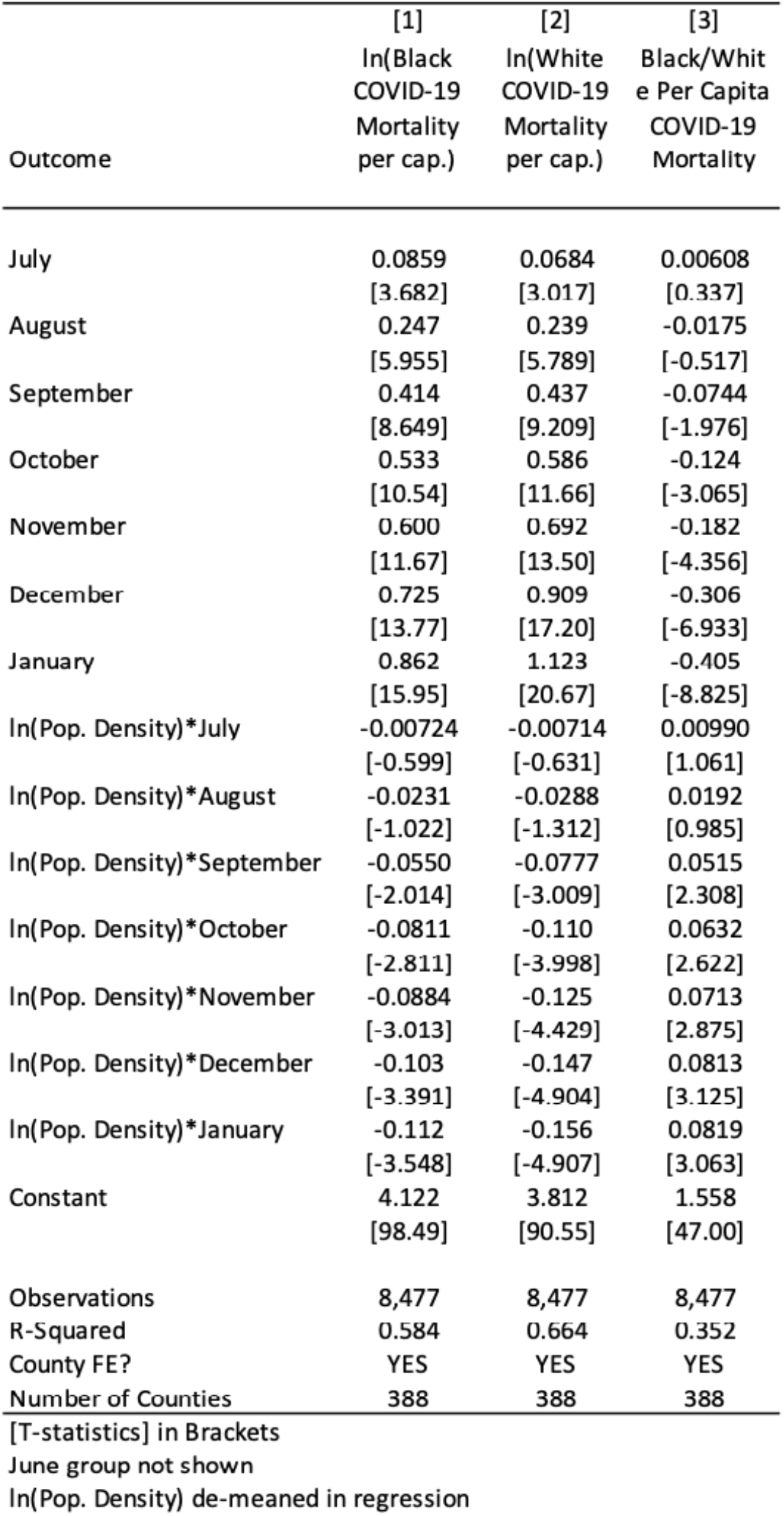
Population Density and Disparity Trends. County-level fixed-effects outcomes for non-Hispanic Black and White per capita mortality, and the mortality ratio, utilizing month indicator variables over time. De-meaned interactions with population density included to estimate heterogeneity by population density.

**Appendix 4:**
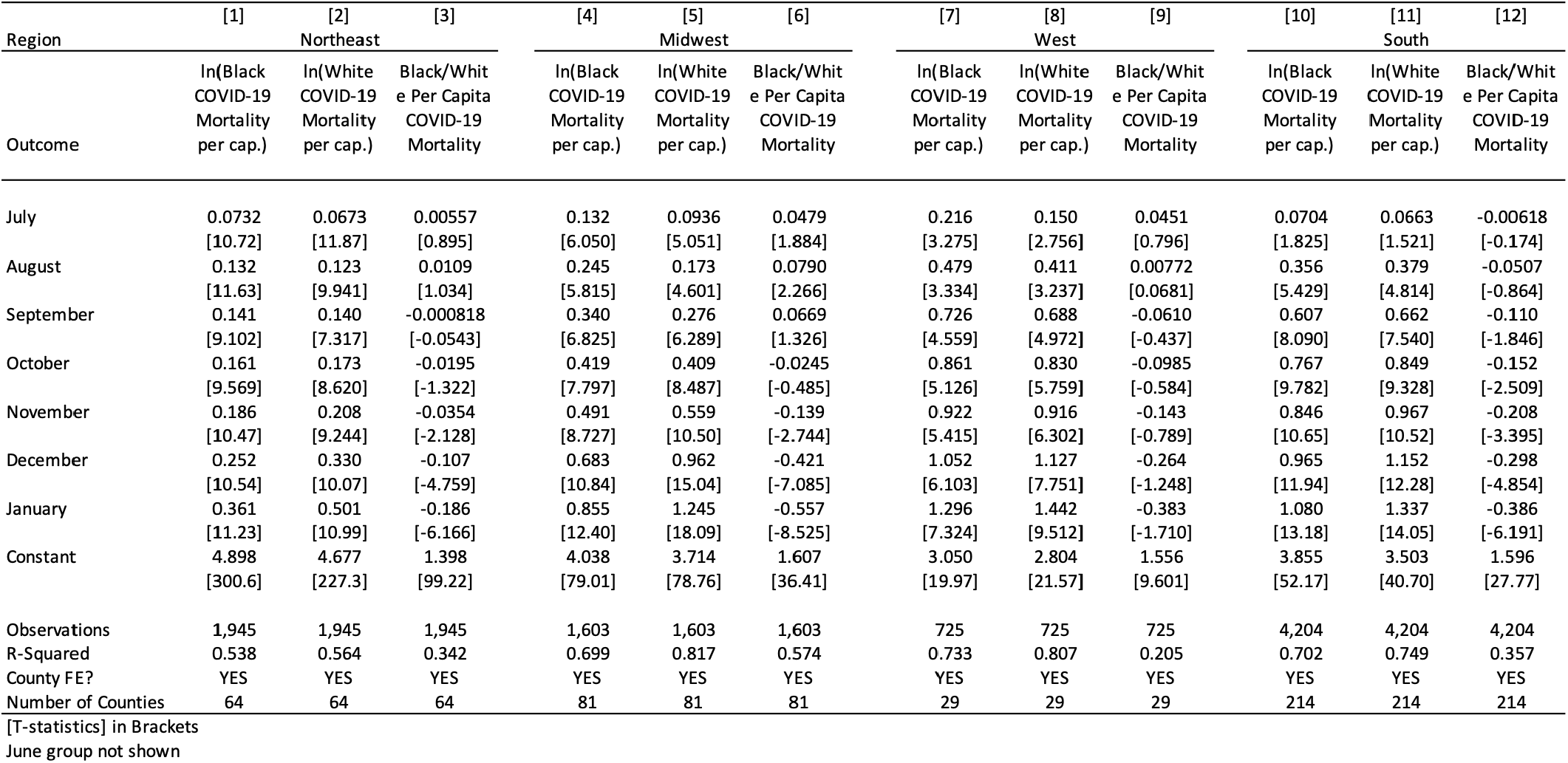
Heterogeneity in COVID Mortality Outcomes by Region. County-level fixed-effects outcomes for non-Hispanic Black and White per capita mortality, and the mortality ratio, utilizing month indicator variables over time, stratified by US Census region.

**Appendix 5:** Heterogeneity in COVID Mortality Outcomes by Region with incidence. County-level fixed-effects outcomes for the Black/White mortality ratio, utilizing month indicator variables over time, interactions with region indicators, and covariates for COVID-19 prevalence.

|  | [1] | [2] |  |  |  |
| --- | --- | --- | --- | --- | --- |
| Outcome | Black/White Per Capita COVID-19 Mortality | Black/White Per Capita COVID-19 Mortality |  |  |  |
| ln(Total Mort. Per Cap.) | -0.183<br>[-4.082] |  |  |  |  |
| ln(Lag Pos. Tests Per Cap.) |  | -0.180<br>[-5.477] |  |  |  |
| July | 0.0105<br>[0.318] | 0.0455<br>[1.372] | October | 0.00536<br>[0.0831] | 0.105<br>[1.629] |
| July*Northeast | 0.00780<br>[0.229] | -0.0259<br>[-0.762] | October*Northeast | 0.00719<br>[0.114] | -0.0594<br>[-0.963] |
| July*Midwest | 0.0576<br>[1.352] | 0.0368<br>[0.865] | October*Midwest | 0.0458<br>[0.603] | 0.0561<br>[0.733] |
| July*West | 0.0701<br>[1.009] | 0.0733<br>[1.032] | October*West | 0.0675<br>[0.364] | 0.0934<br>[0.505] |
| August | 0.0253<br>[0.457] | 0.101<br>[1.750] | November | -0.0317<br>[-0.474] | 0.0781<br>[1.177] |
| August*Northeast | 0.00904<br>[0.164] | -0.0604<br>[-1.059] | November*Northeast | 0.0340<br>[0.523] | -0.0221<br>[-0.355] |
| August*Midwest | 0.0908<br>[1.417] | 0.0609<br>[0.923] | November*Midwest | -0.00939<br>[-0.122] | 0.0196<br>[0.252] |
| August*West | 0.0743<br>[0.560] | 0.0853<br>[0.636] | November*West | 0.0751<br>[0.380] | 0.107<br>[0.548] |
| September | 0.0155<br>[0.258] | 0.109<br>[1.758] | December | -0.0937<br>[-1.324] | 0.0286<br>[0.409] |
| September*Northeast | 0.0102<br>[0.173] | -0.0631<br>[-1.055] | December*Northeast | 0.0450<br>[0.661] | 0.0198<br>[0.311] |
| September*Midwest | 0.106<br>[1.437] | 0.0985<br>[1.306] | December*Midwest | -0.165<br>[-1.945] | -0.110<br>[-1.268] |
| September*West | 0.0672<br>[0.424] | 0.0891<br>[0.562] | December*West | 0.0493<br>[0.219] | 0.0985<br>[0.446] |
|  |  |  | January | -0.152<br>[-2.006] | 0.00127<br>[0.0165] |
|  |  |  | January*Northeast | 0.0545<br>[0.761] | 0.0551<br>[0.828] |
|  |  |  | January*Midwest | -0.195<br>[-2.148] | -0.152<br>[-1.649] |
|  |  |  | January*West | 0.0396<br>[0.166] | 0.0954<br>[0.410] |
|  |  |  | Constant | 1.427<br>[32.75] | 1.316<br>[31.22] |
|  |  |  | Observations | 8,477 | 8,477 |
|  |  |  | R-Squared | 0.431 | 0.436 |
|  |  |  | County FE? | YES | YES |
|  |  |  | Number of Counties | 388 | 388 |
|  |  |  | [T-statistics] in Brackets |  |  |
|  |  |  | June group not shown |  |  |

**Appendix Figure 1:**
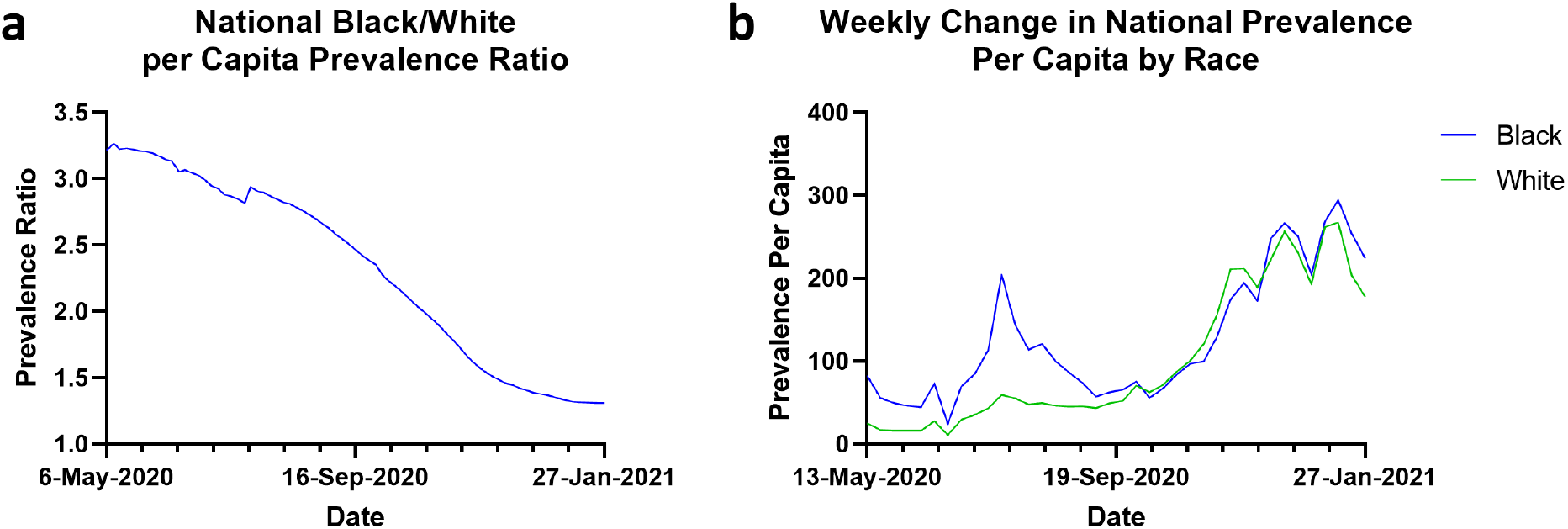
National Trends in COVID-19 Prevalence by Race. (**a**)Weekly Black/White Prevalence Ratio per Capita was calculated by date and subsequently plotted from 5/6/2020 to 1/27/2021. (**b**) Weekly Change in COVID-19 Prevalence by race was calculated by date and subsequently plotted from 5/13/2020 to 1/27/2021.

## Notes

### Competing Interest Statement

E.S.H. is a non-executive Founder of Stratus Medicine, kelaHealth, and MedBlue Data.

### Author Declarations

IRB Approval not applicable. Exempt from Duke IRB Approval.

